# Informing patients that they are at high risk for serious complications of viral infection increases vaccination rates

**DOI:** 10.1101/2021.02.20.21252015

**Authors:** Maheen Shermohammed, Amir Goren, Alon Lanyado, Rachel Yesharim, Donna M. Wolk, Joseph Doyle, Michelle N. Meyer, Christopher F. Chabris

## Abstract

For many vaccine-preventable diseases like influenza, vaccination rates are lower than optimal to achieve community protection. Those at high risk for infection and serious complications are especially advised to be vaccinated to protect themselves. Using influenza as a model, we studied one method of increasing vaccine uptake: informing high-risk patients, identified by a machine learning model, about their risk status. Patients (N=39,717) were evenly randomized to (1) a control condition (exposure only to standard direct mail or patient portal vaccine promotion efforts) or to be told via direct mail, patient portal, and/or SMS that they were (2) at high risk for influenza and its complications if not vaccinated; (3) at high risk according to a review of their medical records; or (4) at high risk according to a computer algorithm analysis of their medical records. Patients in the three treatment conditions were 5.7% more likely to get vaccinated during the 112 days post-intervention (p < .001), and did so 1.4 days earlier (p < .001), on average, than those in the control group. There were no significant differences among risk messages, suggesting that patients are neither especially averse to nor uniquely appreciative of learning their records had been reviewed or that computer algorithms were involved. Similar approaches should be considered for COVID-19 vaccination campaigns.

## Introduction

Controlling the COVID-19 pandemic will require a large majority of people to agree to receive a vaccine. In addition to contributing to population immunity, vaccination is especially important for those at high risk of severe COVID-19 and its complications. Unfortunately, there is a gap between the proportion of people currently interested in being vaccinated and the proportion necessary for effective community protection (1).

The effect of informing patients of their high-risk status on vaccine uptake is not well understood; such information might be motivating (2) or demotivating (3, 4). Moreover, to be scalable, rapid, mass vaccination campaigns targeting high-risk individuals require health data sharing and risk disclosure outside the patient-physician encounter, which patients may view as an invasion of privacy (5). Vaccine encouragement messages that reference a review of patient medical records might therefore backfire.

Finally, it is unknown whether patients will respond differently depending on how their risk status was determined: for example, human versus algorithmic or artificial intelligence-based medical record review. Some laboratory research has documented a pattern of “algorithm aversion,” in which people choose human over superior algorithmic forecasts, especially after they observe the algorithm make an error (6, 7). Yet other laboratory research has found instances of “algorithm appreciation,” in which people followed advice more when they thought it came from algorithms than when they thought it came from humans (8). However, there have been no field experiments directly comparing the effects of disclosing versus not disclosing to individuals the fact that computer algorithms were used to create a specific prediction or rationale for a recommended action. Such experiments would test whether the pro- and anti-algorithm behavior observed in hypothetical lab situations generalizes to real-world, incentive-compatible situations, such as those which will become increasingly common as the use of artificial intelligence and machine learning expands in medical practice (9, 10).

Because annual influenza vaccination campaigns are similar in scale and methods to anticipated COVID-19 vaccination efforts, and because information in a patient’s medical record can be used to predict their level of risk for influenza infection and its complications (11), we addressed these questions by conducting a field experiment during the 2020–21 influenza season with patients predicted by a machine-learning model applied to their electronic health record (EHR) data to be in the top 10% of risk for influenza-related complications.

## Materials and Methods

### Participants

Participants were patients with a primary care provider (PCP) of record at Geisinger (a Pennsylvania health system) who were determined by the Medial EarlySign algorithm to be at high risk for influenza followed by a flu-related complication if they were not vaccinated (see Supplementary Information, SI). The algorithm employed a decision tree-based method (XGBoost) to create a classifier based on 2,371 features related to patient demographics (age, gender, race, etc.), socioeconomic information (years of education, etc.), behavior (smoking, alcohol use, etc.), vital signs, lab test results, diagnoses, medication prescriptions and hospital admissions. The model was trained on data from 2008– 2019 (70%) and tested (20%) and validated (10%) on data from 2012–2019. Risk scores were generated for each patient, and patients with scores in the top 10%, corresponding to a model-predicted probability of getting influenza followed by a complication (if unvaccinated) of at least 11.7%, were enrolled in the study. The Geisinger IRB approved this study and waived the requirement for consent.

### Materials and Procedure

Participants were allocated evenly through parallel random assignment (using R’s “sample” algorithm) to one of four study conditions: to be (1) informed of their high-risk status as determined by a computer algorithm, (2) informed of their high-risk status as determined by a review of their medical records, (3) informed of their high-risk status with method unspecified, or (4) in a control group that was not informed by us of their high-risk status but received all other standard Geisinger messages encouraging influenza vaccination. See SI for the full text of all messages. Notably, at the outset of this flu season, Geisinger contacted all patients with a Geisinger PCP via postcard or the MyChart patient portal with a generic reminder to get vaccinated.

Participants informed of their high-risk status were sent messages via all three of the following channels for which communication was possible: print letters (99% of participants), patient portal (68%), and SMS (55%). Informed participants were also further subdivided based on their risk level: those in the top 3% of evaluated patients (corresponding to a model-predicted risk of complications of at least 25.1%) were told they were in the top 3% of risk; the remaining participants were randomly assigned to be told either that they were in the “top 10% of risk” or at “high risk.”

This study was registered at ClinicalTrials.gov (NCT04323137). Results presented are from an interim analysis conducted to inform rapid COVID-19 vaccination efforts. Reproducible analysis code for this study is available at https://osf.io/8zpyh/. Data will be available in the same repository upon institutional approval.

### Analysis

The primary outcome variable was whether participants were vaccinated between the start of the intervention, September 21, 2020, and January 11, 2021. Vaccination rates were compared between groups using generalized linear models with a binary distribution and log-link function (logistic regression). Assessment of different presentations of risk information (“top 3%”, “top 10%”, and “high”) was conducted within a single logistic regression model, with “top 10%” dummy coded as the reference condition. This allowed comparisons of “top 10%” vs. “high” (to assess if patients with the same risk level are differentially affected by more specific or general risk framing) and of “top 10%” vs. “top 3%” (to assess if patients with similar phrasing are affected by differential risk levels).

We compared the average time to get vaccinated between vaccinated patients in the informed and control groups using Welch’s two-sample t-test (two-sided), with time specified as the number of days since the beginning of the intervention. All statistical analyses were performed in R version 3.2.3 (12).

## Results

Study participants (N = 39,717) had a mean age of 56.4 ± 19.5 (SD) years and were 95.6% white and 65.3% female in a largely rural population in Pennsylvania, U.S.

As shown in Figure 1, patients who were informed of their high-risk status were 5.7% more likely to obtain an influenza vaccination compared with the control group (Odds Ratio: 1.10 [95% CI: 1.05, 1.15], p < .001). Furthermore, among patients who received the vaccination within 112 days of the intervention start, those informed of their risk status (N = 12,679) did so a mean 1.41 days (95% CI: 0.59, 2.23) sooner than those not informed (N = 4,001; t(6554) = 3.38, p < .001, Cohen’s *d* = 0.06.

**Figure 1.**
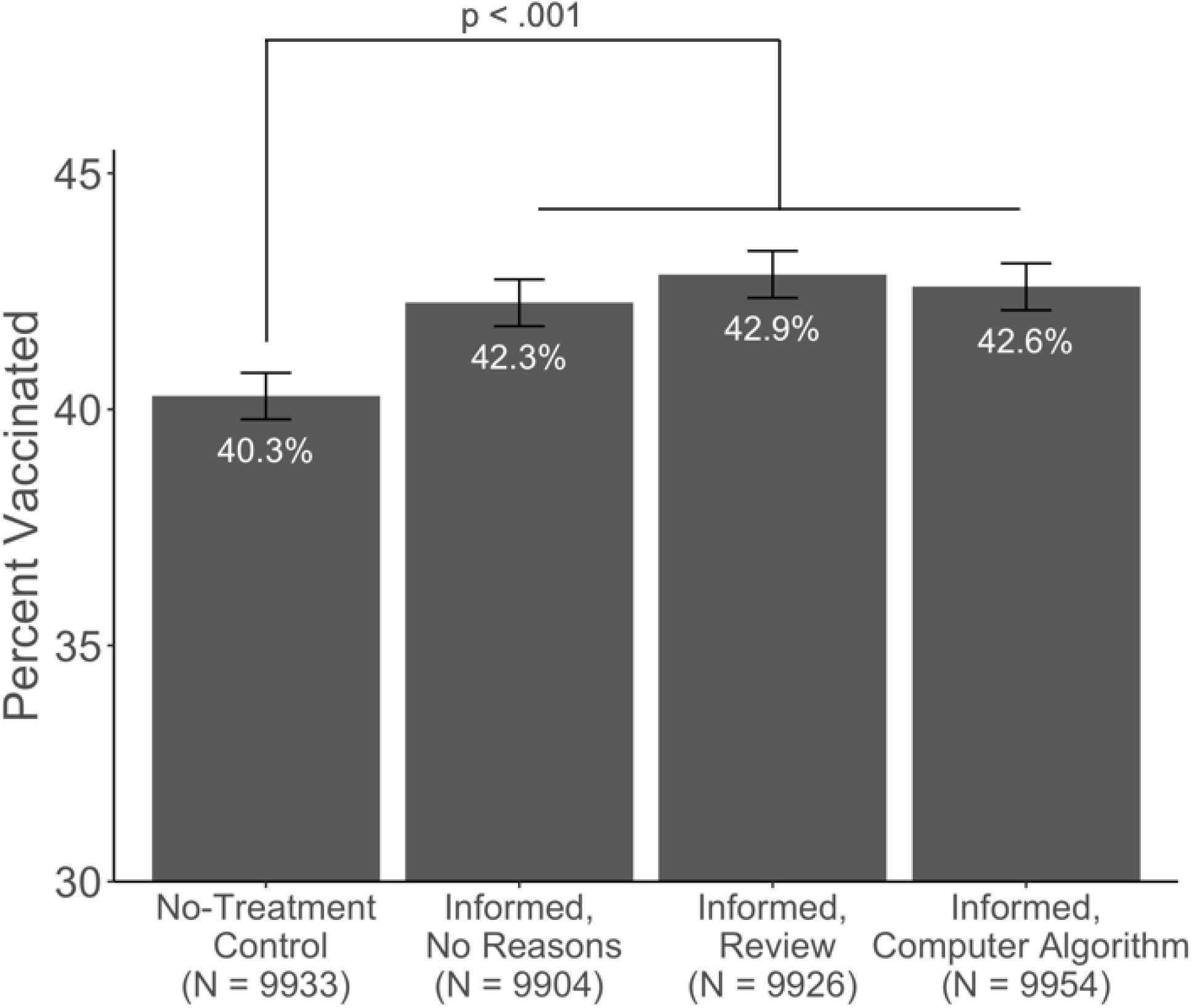
Vaccination rates among patients who were not informed of their high-risk status (“No-Treatment Control”), who were informed of their status without specifying the method with which the status was determined (“Informed, No Reasons”), and who were informed that their status was determined by a review of their medical records (“Informed, Review”) or by a computer algorithm-based analysis of medical records (“Informed, Computer Algorithm”). Error bars represent standard error.

Informing patients that their high-risk status was specifically determined by a “computer algorithm-based analysis of your medical records” performed similarly to simply a “review of your medical records” (OR = 0.99 [95% CI: 0.94, 1.05], p = .710). Patients who received such information about how their status was determined had numerically higher rates of vaccination than those who were simply told they were at high risk; however, these differences were not significant (OR = 1.01 [95% CI: 0.96, 1.07], p = .628 and OR = 1.02 [95% CI: 0.97, 1.08], p = .392 respectively).

Patients under 65 (N = 24,902, 62.7% of the population) were less likely than older patients (N = 14,815) to obtain vaccination (33.1% vs. 57.0%). However, age did not moderate the impact of our interventions. Younger patients exhibited numerically greater relative benefit from being informed of their high-risk status (8.3% vs. 3.2%), but the interaction between age and study condition was not significant (OR = 0.95 [95% CI: 0.87, 1.05], p = .343). There were likewise no significant interactions between age and the method of specifying high risk.

Figure 2 depicts the time course of flu vaccinations before and after our interventions and suggests that intervention impact decreased over time. In line with this, exploratory analysis shows that the effects of the intervention are concentrated in the first month (9/21/20–10/20/20): patients informed of their high-risk status were 9.8% more likely to vaccinate during that time, and 1.3% less likely in the remaining study period (10/21/20–1/11/21).

**Figure 2.**
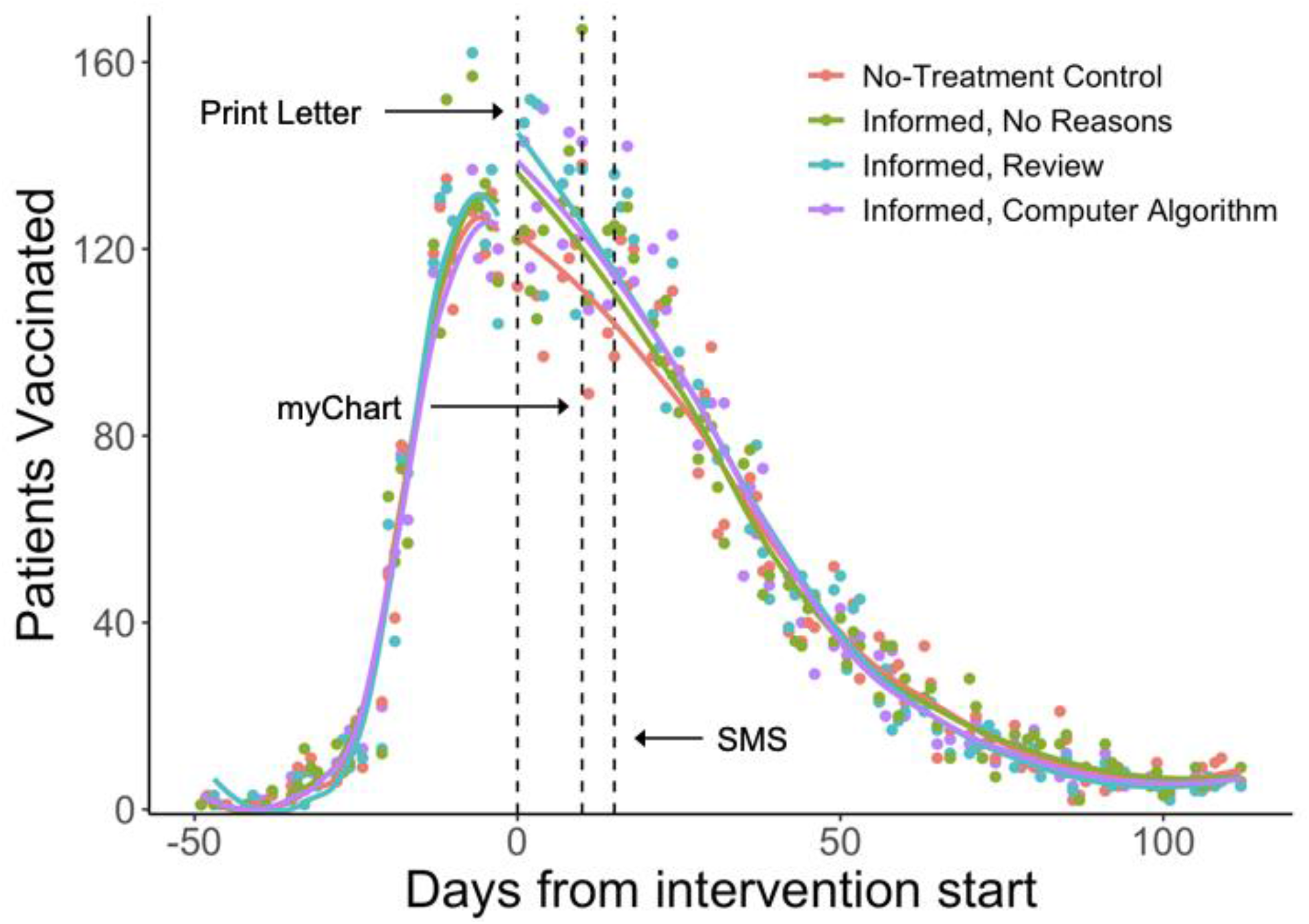
Number of patients vaccinated over time across conditions, excluding weekends and holidays where Geisinger is closed for primary care. Descriptive curves were generated using locally-weighted regression separately for periods before and after start of the intervention. All statistics reported exclude patients vaccinated before the intervention began. Dashed lines indicate the day each communication modality was sent.

The specific risk information that patients received differed by their risk level: those falling in the 97–99th percentiles of risk scores were simply informed that they were in the “top 3% of risk” while the remaining patients (90–96th percentiles) were randomized to be informed that they were either in the “top 10% of risk” or “at high risk” to assess the relative effectiveness of a more specific or general risk description for patients in the same risk stratum. Specificity of risk information (“top 10%” vs. “high”) did not affect vaccinations (OR = 0.99 [95% CI: 0.93, 1.04], p = .595).

## Discussion

We found that informing patients they are at high risk for serious influenza complications increased their likelihood of getting vaccinated. Notably, we found evidence of neither algorithm aversion nor algorithm appreciation, suggesting that disclosure of the use of artificial intelligence, machine learning, and/or algorithms—at least for risk stratification and health communication—may be well received or at least not lead to backlash.

Our intervention resembles many nudges tested in the field: although only modestly effective (13, 14), it is relatively inexpensive, scalable, and preserves choice. Our intervention was also tolerable: Only 3 out of 29,784 patients in the three treatment conditions registered a complaint about the messages they received, and 0.47% of those who received SMS messages replied to opt-out of that specific campaign. Moreover, there are several straightforward opportunities to increase the effectiveness of this intervention, in both the influenza and COVID-19 contexts: They can be: made more timely (e.g., included in reminders for upcoming medical appointments; 15), reinforced with followup messages, and made more personal and convincing by incorporating information about factors contributing to individual recipients’ heightened risk. Health systems increasingly use AI but are unsure whether to disclose this to patients (16). The current findings are expected to generalize well to other to other US health systems with largely rural populations and a diverse range of socioeconomic statuses, but it would be helpful to replicate the study in more racially and ethnically diverse and urban subpopulations.

This work was motivated in part by the hypothesis that interventions that are effective in increasing influenza vaccination may also be effective in the COVID-19 vaccination context, where morbidity and mortality seem to follow a strong risk gradient based on factors such as age, race/ethnicity, and underlying medical conditions. It is true that many individuals considering vaccination will be aware in general terms of their risk for severe COVID-19, but because knowledge of this disease has been evolving continuously and rapidly since the pandemic’s onset, and because media and other public messages about risk factors vary in accuracy and credibility, we believe that directly informing individual patients that they are high risk can be an important tool to nudge fence-sitters to seek or accept vaccination. Moreover, many individuals may be aware of widely publicized risk factors but still not realize they are at high risk. Receiving this personalized information—whether determined by a complex machine learning model or an existing simpler stratification method (17)—from a trusted, local health system or provider (18, 19), combined (as in our study) with information on how they can get vaccinated, could be an especially powerful nudge.

## Data Availability

This study was registered at ClinicalTrials.gov (NCT04323137). Reproducible analysis code for this study is available at https://osf.io/8zpyh/. Data will be available in the same repository upon institutional approval.

## Acknowledgments

We thank Henri Carlo Santos, Ann Marie Tice, Jeffrey Rowe, Rebecca Stametz, William Casey Cauthorn, Carol Hartranft, and Niraj Vishwakarma for their contributions. Research reported in this publication was supported by the National Institute on Aging of the National Institutes of Health under Award Number P30AG034532.

## Notes

**Competing Interest Statement:** A.L. and R.Y. are employed by Medial EarlySign, which designs machine learning-based decision support and risk management tools for health systems. Medial EarlySign employees co-developed the algorithm used to risk-stratify participants but were not involved in the design of this study investigating the effects of disclosing risk status to patients or in the analysis of study data. D.M.W. received grants from Medial EarlySign to support her role in that prior work of co- developing and retrospectively validating the algorithm.

### Competing Interest Statement

A.L. and R.Y. are employed by Medial EarlySign, which designs machine learning-based decision support and risk management tools for health systems. Medial EarlySign employees co-developed the algorithm used to risk-stratify participants but were not involved in the design of this study investigating the effects of disclosing risk status to patients or in the analysis of study data. D.M.W. received grants from Medial EarlySign to support her role in that prior work of co-developing and retrospectively validating the algorithm.

### Clinical Trial

NCT04323137

### Author Declarations

The Geisinger IRB approved this study (IRB # 2020-0290) and waived the requirements for consent (under 45 CFR 46.116(c)-(d)) and for HIPAA authorization (under 45 CFR 164.512(i)(2)(ii)). An NIA-convened Data and Safety Monitoring Board also approved and oversaw this study.

